# Accurate Prediction of Hepatic Disease Severity in MASLD Using a Learned Phenotype

**DOI:** 10.64898/2026.11.14.26368091

**Authors:** Eric V. Strobl, Mark T. Miedel, D. Lansing Taylor, Jaideep Behari

## Abstract

**Background & Aims:** Most prognostic models for metabolic dysfunction-associated steatotic liver disease (MASLD) target prespecified hepatic endpoints, potentially limiting attainable accuracy. We developed and externally validated a model that learns the future hepatic phenotype most reliably predictable from baseline clinical characteristics.

**Methods:** We analyzed 332 HALO-MASLD participants (456 follow-up observations). Sparse Canonical Outcome REgression (SCORE) learned a weighted phenotype from eight hepatic measures, coupled with broken adaptive ridge regression using 99 baseline predictor columns. Participant-level nested five-fold cross-validation compared prediction of the learned phenotype with future liver stiffness measurement (LSM), controlled attenuation parameter (CAP), and FIB-4 benchmarks. Models were externally validated without refitting in the independent NIDDK NAFLD Adult Database 2 cohort (520 participants, 732 follow-up observations). Biological validity and bootstrap stability were also assessed.

**Results:** The learned phenotype emphasized lower platelet count and higher bilirubin, alkaline phosphatase, and LSM. In HALO, predictive correlation was *r* = 0.860 (95% CI [0.824, 0.896]) and predictive *R*^2^ = 0.671, versus *r* = 0.763 for LSM and *r* = 0.469 for CAP predicted from all baseline variables. All six prespecified comparisons remained significant after Holm correction (*p*_Holm_ < 0.05). The phenotype correlated with MRE stiffness (*ρ* = 0.694) and biopsy fibrosis stage (*ρ* = 0.567), and was higher with imaging evidence of portal hypertension (*p* < 0.001). Bootstrap relearning yielded mean phenotype correlation 0.947. In NIDDK, predictive correlation was *r* = 0.836 (95% CI [0.805, 0.863]; predictive *R*^2^ = 0.680), and associations with fibrosis and portal hypertension replicated.

**Conclusions:** Learning the prediction target produced a sparse, clinically interpretable hepatic phenotype more accurately predicted than conventional longitudinal MASLD endpoints in both cohorts. The resulting LIFT-MASLD model is available at https://liftmasld.github.io.

## 1 Impact and Implications

Conventional prognostic models for MASLD target prespecified endpoints, although no single marker fully captures future hepatic disease severity. We instead learned a multivariate hepatic phenotype optimized for longitudinal prediction and identified a sparse composite that was predicted more accurately than future liver stiffness or CAP in both development and external validation cohorts. The learned phenotype was also associated with independent measures of fibrosis and portal hypertension that were not targeted during outcome learning, demonstrating that improved predictability could be achieved while retaining clinical relevance. By forecasting this clinically meaningful hepatic state from a small set of routinely collected base-line measurements, LIFT-MASLD offers clinicians a new approach to longitudinal monitoring and provides researchers with a framework for developing more predictable, biologically meaningful prognostic outcomes.

## 2 Introduction

Metabolic dysfunction-associated steatotic liver disease (MASLD) is the most common chronic liver disease worldwide and spans a broad spectrum from steatosis to steatohepatitis, progressive fibrosis, cirrhosis, and hepatocellular carcinoma (European Association for the Study of the Liver et al., 2024). Approximately one-third of the global adult population has steatotic liver disease, and prevalence continues to increase (Riazi et al., 2022). However, the clinical course of MASLD is highly heterogeneous. Many individuals remain stable for years, whereas others progress to advanced fibrosis, portal hypertension, hepatic decompensation, or liver-related death (Sanyal et al., 2021a). Identifying which patients will develop clinically consequential disease therefore remains a central challenge.

Fibrosis has accordingly become a central focus of MASLD risk stratification because histologic fibrosis stage strongly predicts liver-related morbidity and mortality (Ekstedt et al., 2015; Angulo et al., 2015; Sanyal et al., 2021a). Clinical pathways therefore aim first to identify patients with advanced fibrosis, commonly beginning with the fibrosis-4 index (FIB-4), a blood-based score derived from age, aminotransferases, and platelet count, followed when indicated by imaging-based assessment of liver stiffness (European Association for the Study of the Liver et al., 2024; Shah et al., 2009). Prediction studies have then similarly focused on prespecified fibrosis-related targets, including future progression to fibrosis (Ghandian et al., 2022), liver-stiffness or histologic fibrosis progression (Zhou et al., 2026), and classification of histologic fibrosis stage (Chang et al., 2023). Thus, although predictor models have become increasingly sophisticated, the clinical outcome to be predicted is generally specified in advance.

Fibrosis, however, does not capture the entire hepatic disease state. Clinicians routinely consider other measures such as platelet count, bilirubin, albumin, and evidence of portal hypertension, which reflect complementary aspects of hepatic function and disease severity. Fibrosis measurements are also imperfect: liver biopsy samples only a small fraction of the liver and exhibits substantial sampling variability (Ratziu et al., 2005), while transient elastography measures stiffness as a surrogate for fibrosis and can disagree with histologic staging (Myers et al., 2010). Consequently, a prespecified fibrosis-related endpoint may capture only part of the clinically relevant future disease state and may contain measurement error and biological variability that impose an intrinsic limit on predictive accuracy (Lundberg et al., 2024). Improving longitudinal prediction may therefore require optimizing not only the predictor model but also the clinical state being predicted.

We therefore sought to develop an accurate longitudinal prediction model of future hepatic status in MASLD by jointly optimizing the prediction model and the clinical outcome being predicted, while ensuring that the learned outcome retains a clear clinical interpretation. To this end, we employed Sparse Canonical Outcome REgression (SCORE), which learns an interpretable weighted combination of clinically meaningful outcomes that maximizes prospective predictability from baseline characteristics (Strobl, 2025). We applied SCORE to eight hepatic measures spanning steatosis, liver stiffness, biochemical function and injury, and platelet count, together with sparse predictor selection. We hypothesized that this approach would yield a clinically meaningful hepatic phenotype that could be predicted more accurately and consistently over time than conventional endpoints, while also being associated with independent measures of fibrosis and advanced liver disease. We finally evaluated the generalizability of the trained prediction model in an independent cohort.

## 3 Methods

### 3.1 HALO-MASLD Study

Data were obtained from the University of Pittsburgh Health Assessments and Longitudinal Outcomes in MASLD (HALO-MASLD) study, a prospective longitudinal cohort designed to characterize risk factors and biomarkers associated with MASLD progression. The parent cohort recruited adults aged 18–79 years with diagnosed MASLD through the University of Pittsburgh/UPMC liver program and affiliated research recruitment pathways. Participants underwent detailed annual assessments of hepatic and systemic health, including transient elastography, anthropometric and body-composition measurements, laboratory testing, medication and comorbidity assessment, patient-reported outcomes, lifestyle measures, and biospecimen collection.

For the present study, we included participants with MASLD who had baseline clinical characteristics available at the first study visit and hepatic outcome measurements available at the second and/or third visits. Measurements obtained at the first visit were used exclusively as predictors, whereas measurements obtained at subsequent visits were used to define and evaluate prospective hepatic outcomes.

### 3.2 Ethics Approval

The HALO-MASLD study was approved by the University of Pittsburgh Institutional Review Board (STUDY 21030003). The study was conducted in accordance with the ethical principles of the Declaration of Helsinki, and all participants provided written informed consent before participation. The present analysis was separately reviewed by the University of Pittsburgh IRB and determined not to constitute human subjects research (STUDY26100015).

### 3.3 Study Design and Outcome Definition

We developed a longitudinal clinical prediction model to predict hepatic status at subsequent study visits from clinical characteristics measured at baseline. We restricted the primary analysis to eight prespecified hepatic outcomes chosen to capture complementary dimensions of MASLD severity: controlled attenuation parameter (CAP) for steatosis, FibroScan liver stiffness measurement (LSM) for fibrosis-related stiffness, total bilirubin and albumin for hepatic function, alkaline phosphatase, aspartate aminotransferase (AST), and alanine aminotransferase (ALT) for biochemical injury, and platelet count as a marker associated with advanced fibrosis and portal hypertension. We excluded broader patient-reported measures, such as PROMIS domains, from the primary outcome set because they reflect general physical and psychological health rather than hepatic disease specifically (Cella et al., 2010).

We constructed the predictor matrix exclusively from measurements obtained at the first visit. The resulting matrix contained 99 predictor columns spanning demographic characteristics, anthropometric and body-composition measurements, baseline liver measurements, metabolic laboratory values, comorbidities, medications, lifestyle characteristics, family history, and indicators of missing measurements. We provide the comprehensive list of variables in Appendix Table 4. Sex was defined as sex assigned at birth and encoded as a binary indicator for male sex. We log-transformed selected right-skewed laboratory and elastography variables before analysis. Missing predictor values were median-imputed using medians estimated exclusively within each training sample.

Follow-up observations were included in the primary analysis only when all eight hepatic outcomes were observed; follow-up outcomes were not imputed. We oriented outcomes such that larger values represented greater hepatic disease burden and therefore reversed albumin and platelet count before scaling. Within each training sample, outcomes were min–max scaled and the predictor and outcome matrices were centered separately within each follow-up visit.

### 3.4 Clinical Prediction With SCORE

We used SCORE to determine the multivariate hepatic phenotype that could be most accurately predicted from baseline characteristics. Let **Y**_*t*_ denote the matrix of hepatic outcomes measured at follow-up visit *t* and **X**_*t*_ the corresponding baseline predictor matrix. SCORE learns a nonnegative outcome-weight vector ***α*** together with visit-specific predictor vectors ***β****t* by maximizing predictive dependence between the learned phenotype **Y**_*t*_***α*** and its baseline prediction **X**_*t*_***β***_*t*_ across follow-up visits (Strobl, 2025):

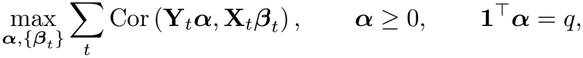

where *q* = 8 was the number of candidate hepatic outcomes.

SCORE alternates between updating the predictor coefficients ***β***_*t*_ conditional on the current outcome weights and updating ***α*** conditional on the current predictor model. To obtain a highly parsimonious clinical prediction model, we replaced the predictor-side ordinary least-squares step with broken adaptive ridge (BAR) regression (Dai et al., 2018). BAR uses iterative reweighted *L*_2_ penalties to approximate *L*_0_ regularization, which penalizes the number of nonzero coefficients rather than continuously shrinking their magnitudes. This typically yields sparser solutions than *L*_1_ regularization while reducing shrinkage of retained predictors. Conditional on ***α***, BAR repeatedly solved weighted ridge regressions of **Y**_*t*_***α*** on **X**_*t*_. At iteration *m*,

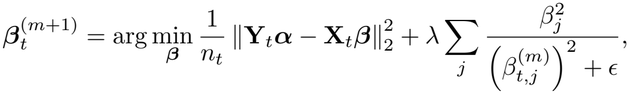

where *λ* denotes the BAR penalty and *ϵ* is a numerical stabilization constant.

We optimized SCORE from multiple initial values to reduce sensitivity to local optima. During cross-validation, we used three starting values for each candidate *λ*. After selecting *λ*, we used 10 starting values for the final fit and retained the solution with the largest SCORE objective.

### 3.5 Nested Cross-Validation

We considered 15 logarithmically spaced values of *λ* between 10^*−*4^ and 10^0^. Predictive performance was estimated using five-fold nested cross-validation with folds defined at the participant level, such that all observations from the same participant remained in either the training or test sample.

Within each outer training fold, we performed five-fold inner cross-validation to select *λ*. The hepatic phenotype and corresponding predictor model were independently learned within each inner training sample, and *λ* was selected to maximize out-of-sample correlation between the observed phenotype and its prediction from baseline characteristics. We then refit SCORE-BAR on the complete outer training sample using the selected *λ* and evaluated the resulting clinical prediction model in the untouched outer test subjects.

After estimating predictive performance, we selected the final BAR penalty by maximizing mean inner-cross-validation performance across the outer training folds and refit SCORE-BAR using all eligible observations. This final fit was used to characterize the learned outcome weights, selected baseline predictors, and associations of the learned phenotype with independent measures of liver disease.

### 3.6 Clinical Prediction Benchmarks

We compared the predictive accuracy of the learned clinical prediction model with models targeting two conventional longitudinal MASLD endpoints: future FibroScan LSM and future CAP. For both endpoints, we first fitted BAR using the complete baseline predictor panel.

For future LSM, we additionally evaluated baseline LSM alone, baseline FIB-4 alone, baseline FIB-4 combined with baseline LSM, and a clinical model containing baseline LSM, age, sex, body mass index, type 2 diabetes, and ALT. The clinical model was fitted using both ordinary linear regression and BAR.

For future CAP, we evaluated baseline CAP alone and a clinical model containing baseline CAP, age, sex, body mass index, type 2 diabetes, and triglycerides, again using both ordinary linear regression and BAR. The same participant-level outer folds were used for all comparisons.

### 3.7 Independent External Validation in NIDDK

We externally validated the trained models using the independent National Institute of Diabetes and Digestive and Kidney Diseases (NIDDK) NAFLD Adult Database 2 cohort (Sanyal et al., 2021b). We used enrollment measurements as baseline predictors and hepatic measurements obtained at approximately 48 and 96 weeks as follow-up outcomes, corresponding to visits 2 and 3 in HALO.

We applied the final SCORE-BAR model trained on the complete HALO cohort to NIDDK without refitting or recalibration. The learned phenotype was calculated using the original HALO outcome weights and min–max scaling parameters. Because albumin, AST, and ALT received zero weights in the final learned phenotype, we required complete follow-up measurements only for the five outcomes with nonzero weights: CAP, LSM, total bilirubin, alkaline phosphatase, and platelet count. The resulting external-validation sample contained 732 follow-up observations. Baseline predictors were harmonized to the HALO definitions, and missing values were imputed using the original HALO training medians.

For comparison, we similarly applied the conventional prediction benchmarks trained on HALO, including the final BAR models for future CAP and LSM using the complete HALO baseline predictor panel. These models retained their original selected predictors, regression coefficients, calibration parameters, and preprocessing transformations. Predictors unavailable in identical form were harmonized where possible; in particular, the HALO cirrhosis diagnosis indicator was approximated using baseline imaging and centrally reviewed histologic evidence of cirrhosis in NIDDK. No model parameters were re-estimated.

### 3.8 Bootstrap Confidence Intervals and Performance Comparisons

We evaluated predictive performance using Pearson correlation, normalized root mean squared error (NRMSE), and predictive *R*^2^. In HALO, Pearson correlations were calculated separately within each outer cross-validation fold and follow-up visit, transformed using Fisher’s *z* transformation, and combined using weights proportional to the corresponding sample size minus three. Prediction errors were normalized by the observed outcome standard deviation within each fold and visit, and NRMSE was calculated by taking the square root of the mean squared standardized errors pooled across all follow-up observations. Predictive *R*^2^ was defined as

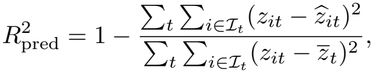

where *z*_*it*_ and 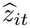 denote the observed and predicted values of the model-specific outcome for participant *i* at follow-up visit *t*, respectively, ℐ_*t*_ denotes the participants observed at visit *t*, and 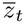 denotes the corresponding observed outcome mean. Thus, predictive *R*^2^ quantified the reduction in squared prediction error relative to visit-specific mean prediction. All three metrics combined observations across visits rather than taking an unweighted average of visit-specific performance. In NIDDK, the same aggregation principles were used, except that correlations and prediction-error normalization were calculated within follow-up visits without cross-validation fold stratification. Each model was evaluated against its designated outcome.

We quantified uncertainty in HALO predictive performance using 2,000 participant-level bootstrap resamples of the out-of-fold predictions generated by nested cross-validation. Participants were resampled so that repeated visits remained together. Predictive performance was recalculated without retraining the models in order to isolate sampling variability in performance evaluation from variability in model estimation (Tsamardinos et al., 2018). We formed 95% confidence intervals from the 2.5th and 97.5th percentiles of the bootstrap distribution. For external validation, we similarly used 2,000 participant-level bootstrap resamples of the NIDDK predictions obtained from the frozen HALO models.

For pairwise comparisons, the same bootstrap sample was applied to both sets of out-of-fold predictions and differences in each performance metric were calculated within each replicate. We used 10,000 participant-level bootstrap resamples for the prespecified comparisons of the learned clinical prediction model with BAR prediction of future LSM using the complete baseline predictor panel and with prediction of future LSM using baseline FIB-4 alone, across all three performance metrics. Two-sided bootstrap *p*-values were calculated by comparing the observed differences with the centered bootstrap distributions, using a finite-sample correction. We selected the former comparator because it used the full set of available baseline predictors while retaining future LSM as the prespecified outcome, and the latter because FIB-4 is widely used as a first-line clinical tool for fibrosis risk stratification in MASLD. The resulting six *p*-values were adjusted using the Holm procedure. The same paired participant-resampling approach was used for comparisons in NIDDK.

### 3.9 Temporal Stability of Predictive Performance

We evaluated persistence of predictive accuracy across follow-up intervals using the signed change in out-of-sample correlation, Δ = *r*_visit 3_ *− r*_visit 2_. Values near zero indicated stable predictive performance, whereas negative values indicated lower predictive accuracy at visit 3. We obtained 95% confidence intervals for visit-specific correlations and their changes using 2,000 participant-level bootstrap resamples in both HALO and NIDDK.

### 3.10 Validation Against Independent Measures of Liver Disease

We evaluated the clinical interpretation of the learned hepatic phenotype using complementary measures of liver disease severity available in subsets of participants. We prioritized MRE liver stiffness and histologic fibrosis stage as established measures of fibrotic disease, and imaging evidence of portal hypertension as a clinically consequential manifestation of advanced liver disease. Each measure was matched to the corresponding follow-up visit. Associations with continuous or ordinal measures were quantified using Spearman rank correlation to allow for monotonic but potentially nonlinear relationships. For binary imaging evidence of portal hypertension, we compared mean learned phenotype values between participants with and without portal hypertension. We obtained 95% confidence intervals and two-sided *p*-values using 10,000 participant-level bootstrap resamples, with *p*-values calculated from centered bootstrap distributions.

We repeated these analyses in NIDDK using centrally reviewed histologic fibrosis stage and imaging evidence of portal hypertension. Histologic fibrosis stages were harmonized to a common ordinal scale, and imaging evidence of portal hypertension was defined by documented splenomegaly, ascites, or other portal-hypertension features on abdominal ultrasound, CT, or MRI. These measures were matched to the corresponding follow-up visit. The same participant-level bootstrap procedures were applied. Quantitative MRE stiffness measurements were unavailable in NIDDK.

### 3.11 Bootstrap Stability of the Learned Phenotype

We assessed reproducibility of the model-defined phenotype using 200 participant-level bootstrap samples. For each replicate, we resampled participants with replacement and repeated five-fold cross-validation over the original 15-value *λ* grid using three SCORE starting values for each candidate *λ*. We then relearned the first hepatic phenotype using the selected *λ* and 10 starting values, thereby incorporating variability arising from participant sampling, BAR tuning, and SCORE optimization.

We quantified stability of the outcome weights using cosine similarity between the bootstrap estimate 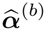 and the original estimate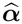. We separately quantified stability of the resulting phenotype using a common reference outcome matrix **Y**_ref_, containing the pooled follow-up observations after outcome orientation and min–max scaling, without visit centering. For each bootstrap replicate, we calculated the correlation between 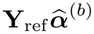 and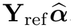. Both quantities were summarized using their means and 2.5th and 97.5th percentiles.

## 4 Results

### 4.1 Cohort Characteristics

The HALO cohort included 332 participants contributing 456 follow-up observations, comprising 297 observations at visit 2 and 159 at visit 3; 124 participants contributed observations at both follow-up visits. Baseline characteristics are summarized in Table 1.

**Table 1:** Characteristics of the HALO cohort. Values are mean (SD) for age and BMI, median [interquartile range] for hepatic measures, and *n* (%) for categorical variables.

| **Characteristic** | **Available *n*** | **Value** |
|---|---|---|
| *Demographic and metabolic characteristics* |  |  |
| Age, years | 332 | 56.3 (13.1) |
| Male sex | 332 | 118 (35.5%) |
| White race | 332 | 307 (92.5%) |
| Black race | 332 | 16 (4.8%) |
| Asian race | 332 | 9 (2.7%) |
| Hispanic ethnicity | 332 | 7 (2.1%) |
| BMI, kg/m<sup>2</sup> | 330 | 34.3 (7.8) |
| Obesity | 332 | 250 (75.3%) |
| Type 2 diabetes | 332 | 137 (41.3%) |
| Hypertension | 332 | 214 (64.5%) |
| Dyslipidemia | 332 | 222 (66.9%) |
| *Baseline hepatic characteristics* |  |  |
| CAP, dB/m | 325 | 301 [262–328] |
| Liver stiffness, kPa | 325 | 6.8 [5.0–10.0] |
| FIB-4 | 310 | 1.10 [0.79–1.58] |
| Total bilirubin, mg/dL | 317 | 0.60 [0.50–0.80] |
| Albumin, g/dL | 317 | 4.3 [4.1–4.6] |
| Alkaline phosphatase, U/L | 317 | 69 [58–86] |
| AST, U/L | 318 | 25 [19–36] |
| ALT, U/L | 318 | 31 [20–47.8] |
| Platelets, 10<sup>9</sup>/L | 314 | 241 [197–281] |

The cohort was predominantly White and had a high burden of metabolic disease. Baseline liver disease was heterogeneous: CAP indicated substantial steatosis, whereas liver stiffness and FIB-4 suggested that most participants did not have advanced fibrosis. Liver synthetic function and platelet counts were generally preserved, although the distribution of liver stiffness indicated a subgroup with more advanced disease.

### 4.2 SCORE-BAR Identified a Sparse Hepatic Phenotype

SCORE-BAR identified the sparse hepatic phenotype plotted in Figure 1A. Reverse-coded platelet count accounted for 44.9% of the normalized outcome weight, followed by total bilirubin (20.8%), alkaline phosphatase (17.6%), FibroScan liver stiffness (15.9%), and CAP (0.8%). Albumin, AST, and ALT received zero weight. The predicted target therefore emphasized thrombocytopenia, biochemical evidence of hepatic dysfunction or cholestasis, and liver stiffness rather than steatosis or aminotransferase elevation.

**Figure 1:**
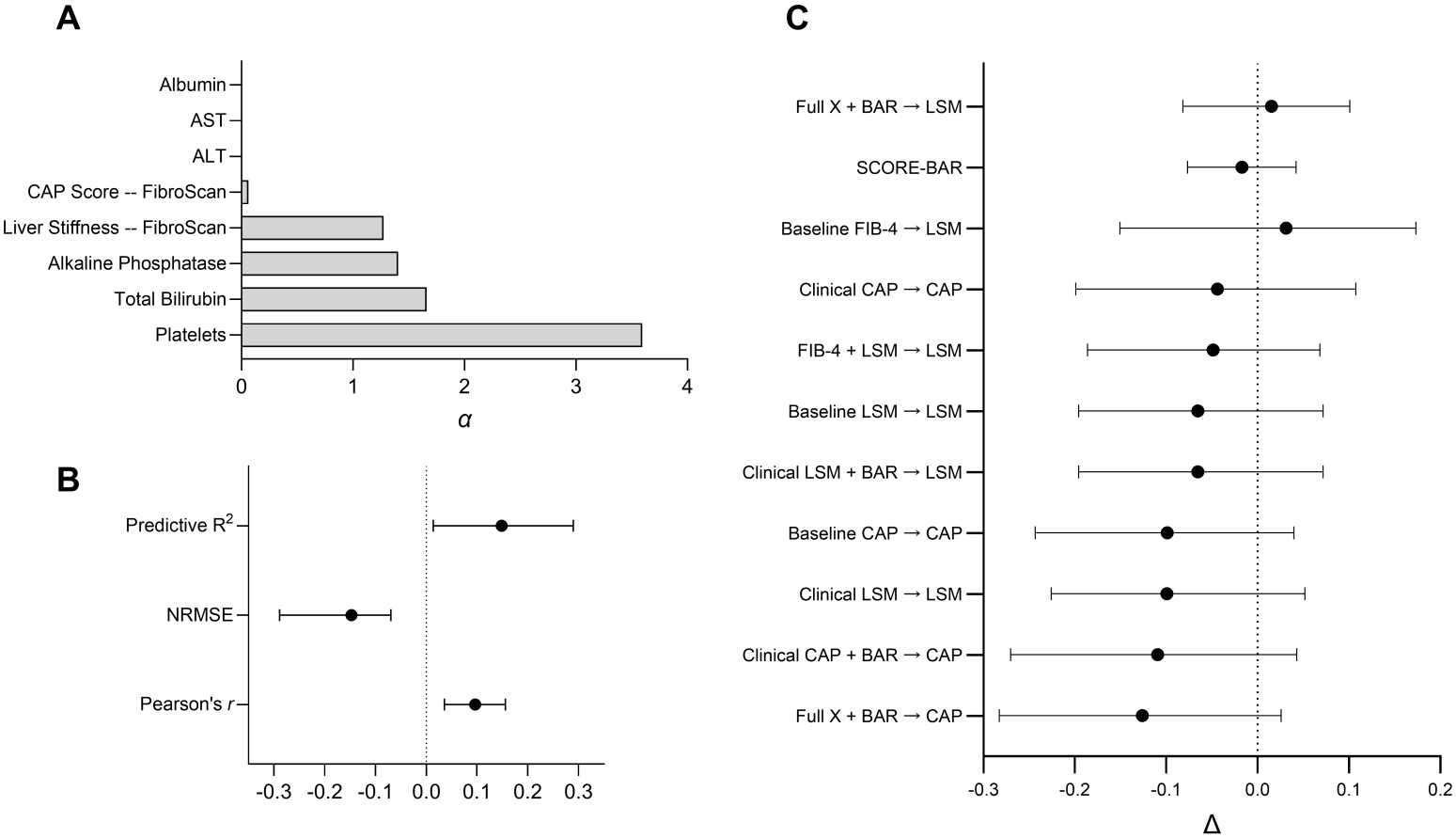
Learned hepatic phenotype and predictive performance. (A) SCORE-BAR outcome weights. (B) Differences versus BAR prediction of future LSM using all baseline predictors: Pearson *r* (+0.096, *p* = 0.002), NRMSE (*−*0.147, *p* = 0.021), and predictive *R*^2^ (+0.149, *p* = 0.036). *p*-values are from two-sided paired participant-level bootstrap tests (10,000 resamples). (C) Changes in predictive correlation from visit 2 to visit 3. Points indicate estimates, horizontal bars indicate 95% bootstrap confidence intervals, and dotted lines indicate zero differences. All visit-change confidence intervals included zero.

The corresponding BAR predictor was also highly sparse. Of the 99 available baseline predictor columns, only six had nonzero coefficients at either follow-up visit. Four represented measured baseline characteristics— liver stiffness, bilirubin, alkaline phosphatase, and platelet count—whereas two represented missingness indicators. Thus, prediction of the learned hepatic phenotype ultimately relied on a small subset of the available baseline information.

### 4.3 SCORE-BAR Achieved the Highest Predictive Accuracy

SCORE-BAR achieved the highest predictive accuracy across the evaluated targets and models (Table 2). Its overall nested cross-validated correlation was *r* = 0.860 and predictive *R*^2^ = 0.671, compared with *r* = 0.763 and *R*^2^ = 0.523 for future LSM, and *r* = 0.469 and *R*^2^ = 0.176 for future CAP, when each was predicted using the complete baseline predictor panel with BAR.

**Table 2:** Nested cross-validated predictive performance. Values are point estimates with participant-level bootstrap 95% confidence intervals [lower bound, upper bound]. *X* denotes the complete set of 99 baseline predictor columns. SCORE-BAR achieved the highest Pearson correlation and predictive *R*^2^, and the lowest NRMSE.

| **Model** | **Pearson *r*** | **NRMSE** | **Predictive *R*<sup>2</sup>** |
|---|---|---|---|
| Baseline CAP → CAP | 0.477 [0.396, 0.565] | 0.897 [0.861, 0.966] | 0.196 [0.102, 0.265] |
| Clinical CAP → CAP | 0.455 [0.377, 0.544] | 0.911 [0.873, 0.986] | 0.163 [0.059, 0.247] |
| Clinical CAP + BAR → CAP | 0.448 [0.364, 0.538] | 0.914 [0.872, 0.990] | 0.160 [0.049, 0.246] |
| Full *X* + BAR → CAP | 0.469 [0.388, 0.558] | 0.908 [0.871, 0.981] | 0.176 [0.078, 0.252] |
| Baseline FIB-4 → LSM | 0.552 [0.401, 0.663] | 0.849 [0.789, 1.011] | 0.259 [0.105, 0.380] |
| Baseline LSM → LSM | 0.730 [0.666, 0.824] | 0.802 [0.717, 0.997] | 0.367 [0.193, 0.492] |
| FIB-4 + LSM → LSM | 0.747 [0.677, 0.823] | 0.752 [0.684, 0.920] | 0.430 [0.285, 0.547] |
| Clinical LSM → LSM | 0.721 [0.658, 0.811] | 0.795 [0.712, 0.994] | 0.397 [0.233, 0.512] |
| Clinical LSM + BAR → LSM | 0.730 [0.666, 0.824] | 0.802 [0.717, 0.997] | 0.367 [0.193, 0.492] |
| Full *X* + BAR → LSM | 0.763 [0.707, 0.826] | 0.683 [0.636, 0.837] | 0.523 [0.407, 0.606] |
| SCORE-BAR | **0.860 [0.824, 0.896]** | **0.536 [0.484, 0.623]** | **0.671 [0.555, 0.765]** |

Paired participant-level bootstrap comparisons confirmed that the learned hepatic phenotype was more predictable than future LSM predicted using the complete baseline predictor panel with BAR (Figure 1B). Predictive correlation was higher by 0.096 (95% CI [0.036, 0.156]; *p* = 0.002), NRMSE was lower by 0.147 (95% CI [0.069, 0.289]; *p* = 0.021), and predictive *R*^2^ was higher by 0.149 (95% CI [0.014, 0.290]; *p* = 0.036).

Differences were also larger relative to baseline FIB-4 prediction of future LSM, with predictive correlation higher by 0.308 (95% CI [0.208, 0.447]; *p <* 0.001), NRMSE lower by 0.313 (95% CI [0.237, 0.458]; *p* = 0.001), and predictive *R*^2^ higher by 0.413 (95% CI [0.270, 0.571]; *p <* 0.001). All six comparisons remained significant after Holm correction (*p*_Holm_ *≤* 0.042).

### 4.4 Predictive Accuracy Was Maintained Across Follow-Up Visits

SCORE-BAR remained stable across the two follow-up visits (Figure 1C), with correlations of *r* = 0.865 at visit 2 and *r* = 0.848 at visit 3 (Δ = *−*0.017, 95% CI [-0.077, 0.042]). BAR prediction of LSM using the complete baseline predictor panel was similarly stable (Δ = 0.015, 95% CI [-0.082, 0.101]), whereas the absolute changes among the other LSM models ranged from 0.031 to 0.099.

CAP prediction generally showed greater deterioration in point estimates over time. Baseline CAP decreased from *r* = 0.510 to *r* = 0.411 (Δ = *−* 0.099, 95% CI [-0.243, 0.040]), while BAR prediction of CAP using the complete baseline predictor panel decreased from *r* = 0.510 to *r* = 0.384 (Δ = *−* 0.126, 95% CI [-0.283, 0.026]). However, all confidence intervals included zero, indicating that the observed changes were not statistically distinguishable from zero. Thus, SCORE-BAR combined high absolute predictive accuracy with little observed change in performance across follow-up intervals.

### 4.5 The Learned Phenotype Was Associated With Fibrosis and Portal Hypertension

Independent validation supported an association between the learned phenotype and fibrosis severity. The strongest associations were observed with MRE liver stiffness (*ρ* = 0.694, 95% CI [0.477, 0.827], *p <* 0.001; Figure 2A) and histologic fibrosis stage (*ρ* = 0.567, 95% CI [0.340, 0.726], *p <* 0.001; Figure 2B). The phenotype was also significantly higher among patients with imaging evidence of portal hypertension than among those without (mean difference = 0.127, 95% CI [0.085, 0.170], *p <* 0.001; Figure 2C). Together, these associations supported the construct validity of the learned composite outcome as a measure of clinically meaningful advanced liver disease, despite fibrosis severity and portal hypertension not being specified as targets during outcome learning.

**Figure 2:**
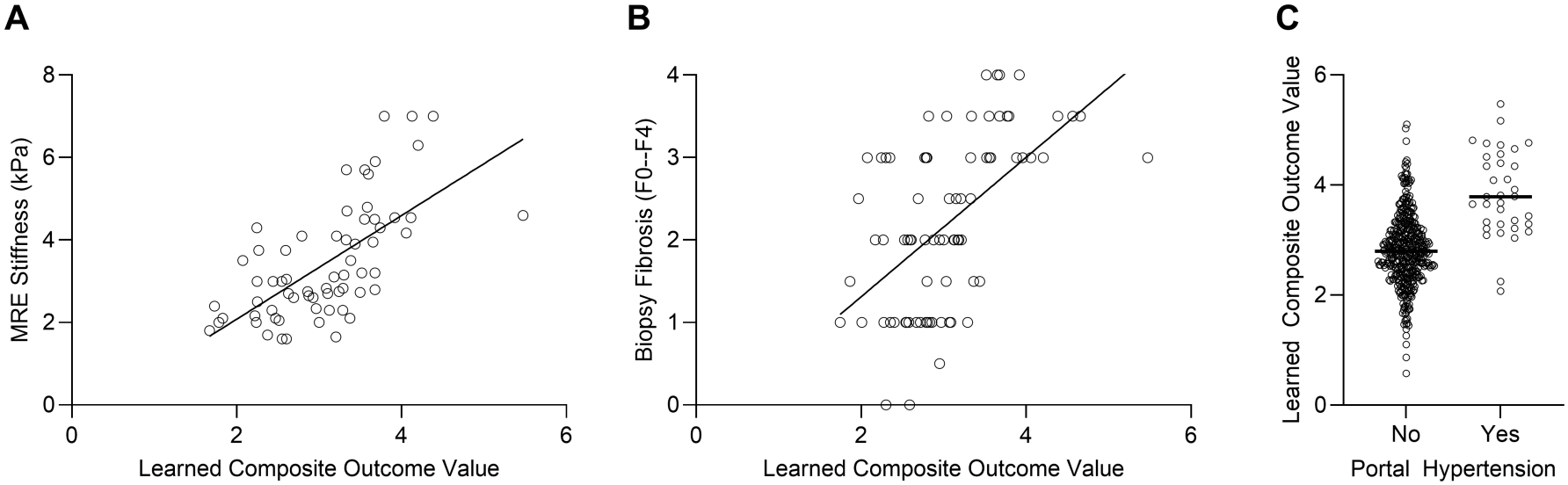
Association of the learned hepatic phenotype with independent measures of liver disease. (A) MRE liver stiffness (Spearman *ρ* = 0.694, *p <* 0.001). (B) Histologic fibrosis stage (Spearman *ρ* = 0.567, *p <* 0.001). (C) Imaging evidence of portal hypertension (mean composite difference = 1.017, *p <* 0.001). All *p*-values are two-sided and were obtained using 10,000 participant-level bootstrap resamples.

### 4.6 The Learned Phenotype Was Reproducible Under Resampling and Retuning

The hepatic phenotype remained stable when the complete SCORE-BAR procedure was repeated across participant-level bootstrap samples, including reselection of the BAR penalty. The mean cosine similarity between the original and bootstrap-relearned outcome-weight vectors was 0.901 (95% percentile interval 0.738–0.996).

Stability was even greater at the phenotype level. The mean correlation between the original learned phenotype and phenotypes constructed using bootstrap-relearned outcome weights was 0.947 (95% percentile interval 0.850-0.998). Thus, variation in the precise outcome weights rarely produced substantial changes in the resulting patient-level phenotype.

### 4.7 External Validation Replicated the Predictive Accuracy and Biological Relevance of SCORE-BAR

In the independent NIDDK NAFLD Adult Database 2 cohort, SCORE-BAR again achieved the highest predictive accuracy among the evaluated models (Table 3), with a Pearson correlation of *r* = 0.836, NRMSE of 0.563, and predictive *R*^2^ = 0.680 across 732 follow-up observations. Predictive accuracy remained high across follow-up visits, although correlation decreased from *r* = 0.863 at visit 2 to *r* = 0.792 at visit 3 (Δ = *−* 0.070, 95% CI [-0.119, -0.028]). Independent biological validation further demonstrated that the learned phenotype correlated with histologic fibrosis stage (*ρ* = 0.358, 95% CI [0.069, 0.600], *p* = 0.008) and was significantly higher among patients with imaging evidence of portal hypertension than among those without (mean difference = 0.962, 95% CI [0.496, 1.432], *p <* 0.001). MRE stiffness measurements were unavailable in NIDDK, precluding external replication of the corresponding HALO association. Together, these findings support the generalizability of SCORE-BAR’s predictive performance and the biological relevance of its learned phenotype to fibrosis-related liver disease severity.

**Table 3:**
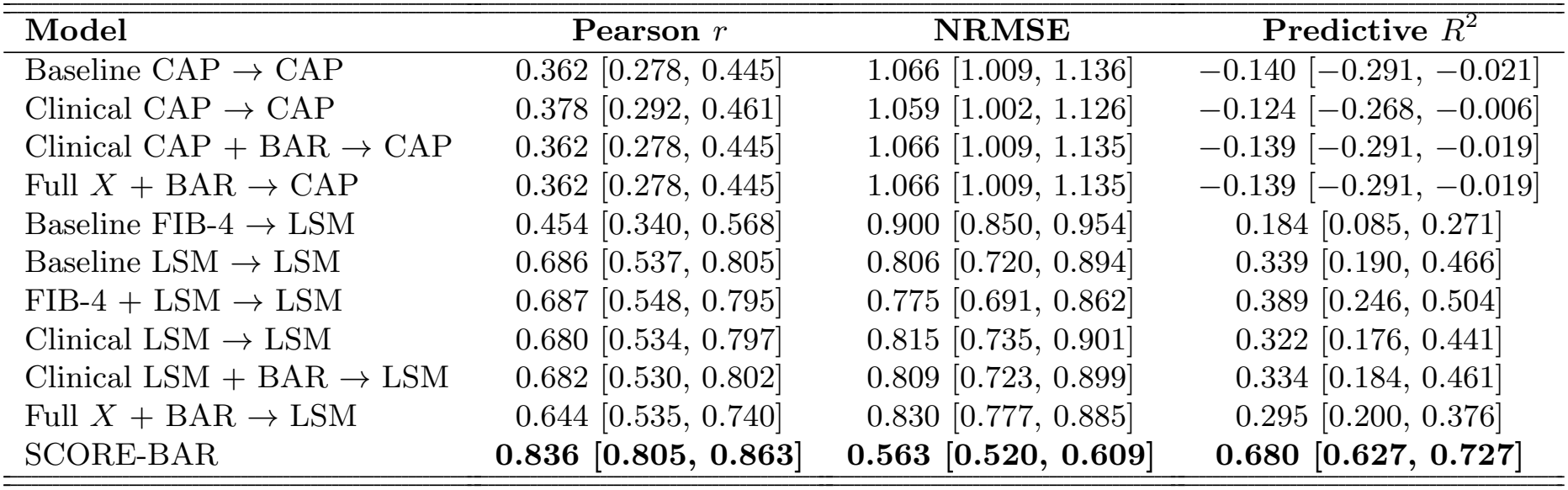
External validation of longitudinal hepatic prediction in the NIDDK NAFLD Adult Database 2 cohort. The analysis included 520 participants contributing 732 follow-up observations. Values are point estimates with 95% bootstrap confidence intervals.

**Table 4:**
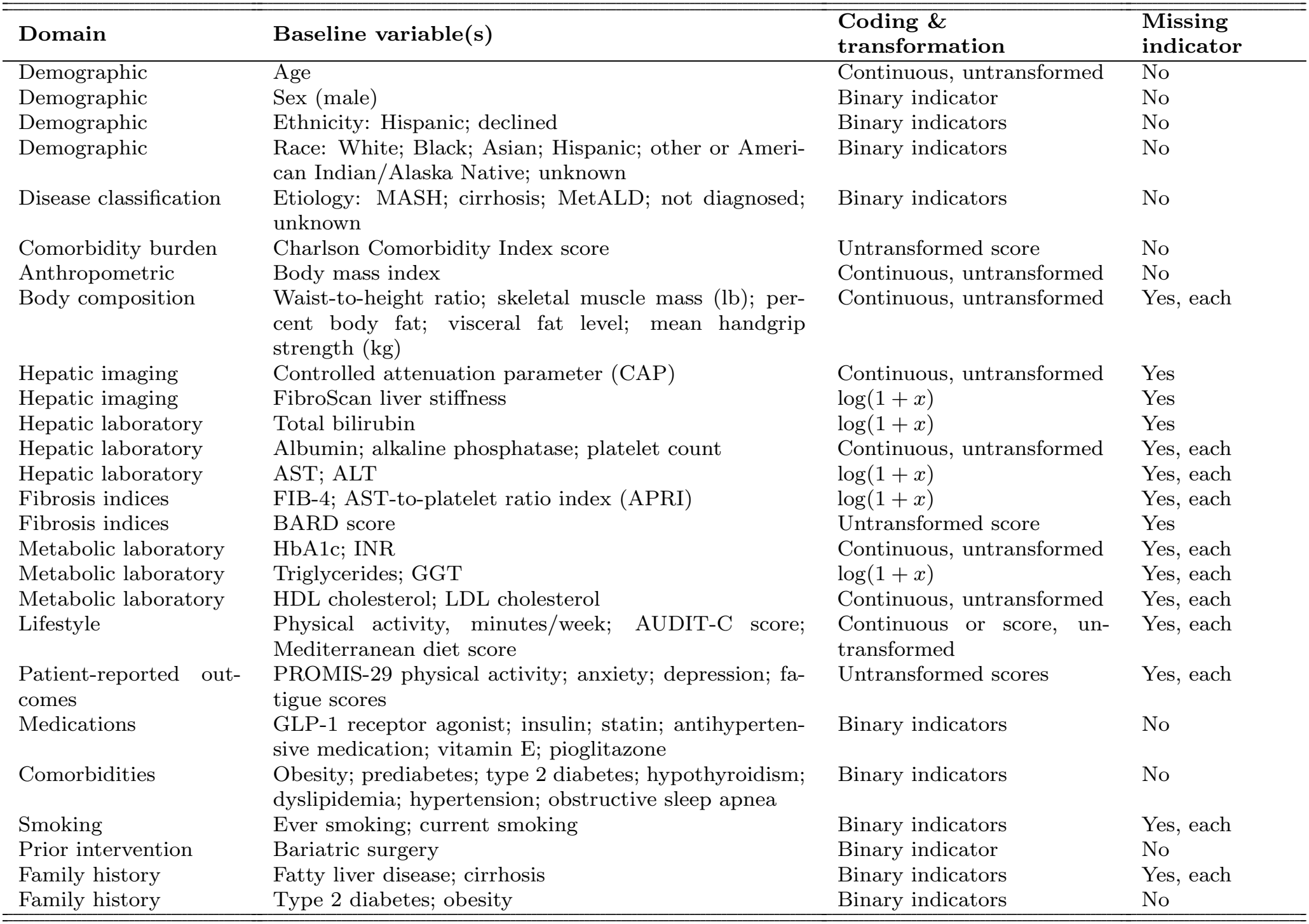
Baseline variables included in the clinical predictor matrix. Sixty-six substantive variables generated 99 predictor columns after addition of 33 variable-specific missingness indicators.

| **Domain** | **Baseline variable(s)** | **Coding & transformation** | **Missing indicator** |
|---|---|---|---|
| Demographic | Age | Continuous, untransformed | No |
| Demographic | Sex (male) | Binary indicator | No |
| Demographic | Ethnicity: Hispanic; declined | Binary indicators | No |
| Demographic | Race: White; Black; Asian; Hispanic; other or Ameri-can Indian/Alaska Native; unknown | Binary indicators | No |
| Disease classification | Etiology: MASH; cirrhosis; MetALD; not diagnosed; unknown | Binary indicators | No |
| Comorbidity burden | Charlson Comorbidity Index score | Untransformed score | No |
| Anthropometric | Body mass index | Continuous, untransformed | No |
| Body composition | Waist-to-height ratio; skeletal muscle mass (lb); per-cent body fat; visceral fat level; mean handgrip strength (kg) | Continuous, untransformed | Yes, each |
| Hepatic imaging | Controlled attenuation parameter (CAP) | Continuous, untransformed | Yes |
| Hepatic imaging | FibroScan liver stiffness | log(1 + *x*) | Yes |
| Hepatic laboratory | Total bilirubin | log(1 + *x*) | Yes |
| Hepatic laboratory | Albumin; alkaline phosphatase; platelet count | Continuous, untransformed | Yes, each |
| Hepatic laboratory | AST; ALT | log(1 + *x*) | Yes, each |
| Fibrosis indices | FIB-4; AST-to-platelet ratio index (APRI) | log(1 + *x*) | Yes, each |
| Fibrosis indices | BARD score | Untransformed score | Yes |
| Metabolic laboratory | HbA1c; INR | Continuous, untransformed | Yes, each |
| Metabolic laboratory | Triglycerides; GGT | log(1 + *x*) | Yes, each |
| Metabolic laboratory | HDL cholesterol; LDL cholesterol | Continuous, untransformed | Yes, each |
| Lifestyle | Physical activity, minutes/week; AUDIT-C score; Mediterranean diet score | Continuous or score, un-transformed | Yes, each |
| Patient-reported out-comes | PROMIS-29 physical activity; anxiety; depression; fa-tigue scores | Untransformed scores | Yes, each |
| Medications | GLP-1 receptor agonist; insulin; statin; antihyperten-sive medication; vitamin E; pioglitazone | Binary indicators | No |
| Comorbidities | Obesity; prediabetes; type 2 diabetes; hypothyroidism; dyslipidemia; hypertension; obstructive sleep apnea | Binary indicators | No |
| Smoking | Ever smoking; current smoking | Binary indicators | Yes, each |
| Prior intervention | Bariatric surgery | Binary indicator | No |
| Family history | Fatty liver disease; cirrhosis | Binary indicators | Yes, each |
| Family history | Type 2 diabetes; obesity | Binary indicators | No |

## 5 Discussion

We developed and both internally and externally validated a sparse clinical prediction model for future hepatic status in MASLD. Rather than prespecifying a single longitudinal marker as the prediction target, the model used SCORE to identify the multivariate hepatic phenotype that could be predicted most reliably from baseline clinical characteristics. The resulting phenotype emphasized lower platelet count, higher bilirubin, higher alkaline phosphatase, and greater liver stiffness and was predicted more accurately than conventional future LSM or CAP in both the HALO and NIDDK cohorts. Predictive accuracy remained high across follow-up visits, while independent biological validation in both cohorts supported the construct validity of the learned phenotype as a clinically meaningful measure of advanced liver disease.

The results suggest that the choice of prediction target can be as important as the choice of prediction algorithm. Most clinical prognostic models prespecify a marker such as liver stiffness or fibrosis stage and then seek to improve performance by adding predictors or increasing model flexibility. Prediction, however, remains constrained by the intrinsic predictability and measurement properties of that target. In the present study, allowing the clinical prediction model to learn a multivariate target produced a future hepatic state that could be predicted substantially more accurately from baseline information. This strategy is consistent with task-aligned outcome learning, in which the most useful outcome representation may depend on the inferential goal (Strobl, 2026). It does not replace conventional fibrosis endpoints when fibrosis itself is the clinical question, but offers a complementary strategy when the objective is accurate prospective prediction of overall hepatic disease severity.

The phenotype learned by the model was clinically coherent. Platelet count received the greatest weight, followed by bilirubin, alkaline phosphatase, and LSM, whereas CAP, AST, and ALT received little or no weight. Its associations with MRE liver stiffness, histologic fibrosis stage, and evidence of portal hypertension supported the construct validity of the learned composite outcome as a clinically meaningful representation of advanced liver disease, even though these measures were not explicitly targeted during outcome learning. This interpretation was further supported by independent replication in NIDDK, where the phenotype was associated with centrally reviewed histologic fibrosis stage and was significantly higher among patients with imaging evidence of portal hypertension. The predictor model was also highly sparse: only six of the 99 baseline predictor columns received nonzero coefficients, requiring just five routinely collected clinical measurements. Thus, the model provides interpretability on both sides of the prediction task: what clinical state is being predicted and which baseline measurements are used to predict it. We refer to the final prediction model as LIFT-MASLD (Liver Integrated Forecasting Tool for MASLD) and have implemented it as a web-based calculator at https://liftmasld.github.io.

These properties suggest potential applications in clinical monitoring and precision medicine. For example, a patient with modest baseline liver stiffness but low platelet count and elevated bilirubin might have a relatively high predicted LIFT-MASLD value, indicating greater anticipated hepatic disease burden than liver stiffness alone would suggest. Such predictions could complement conventional assessments when monitoring patients longitudinally. More broadly, LIFT-MASLD could be incorporated into a patient digital twin framework integrating computational predictions with experimental measurements from patient-derived liver microphysiological systems (Miedel et al., 2025). Comparing predicted hepatic status with disease characteristics and drug responses measured in patient-derived liver MPS could help connect clinical forecasts to underlying biological mechanisms and support the development of personalized therapeutic strategies.

Several limitations should be considered. First, the prediction model uses a linear predictor structure; nonlinear approaches could potentially capture additional interactions, although the present formulation offers interpretability and sparsity. Second, the analysis included only two subsequent follow-up visits, and longer follow-up will be needed to determine whether the model-defined phenotype correlates with other clinical outcomes such as hepatic decompensation, hepatocellular carcinoma, transplantation, or liver-related mortality. Third, although the model was externally validated in the independent NIDDK cohort, HALO was a single-center cohort that was predominantly White, and additional validation across diverse populations and clinical settings would further establish transportability. Finally, the predictors were primarily routinely available clinical, laboratory, anthropometric, and imaging variables; molecular measurements may provide additional predictive information.

In conclusion, we developed and externally validated a sparse clinical prediction model for longitudinal hepatic status in MASLD. By allowing the prediction target itself to be learned, the model identified a clinically meaningful hepatic phenotype with independent evidence of construct validity. This phenotype was more accurately predicted than conventional future LSM or CAP in both cohorts, using only a small set of routinely collected baseline clinical measurements.

## Data Availability

HALO-MASLD data are not publicly available due to institutional and participant privacy restrictions. Requests for access may be directed to Dr. Jaideep Behari and are subject to institutional review and approval. NIDDK NAFLD Adult Database 2 data are available through the NIDDK Central Repository.

https://repository.niddk.nih.gov/study/88

## Abbreviations

ALT: alanine aminotransferase
APRI: AST-to-platelet ratio index
AST: aspartate aminotransferase
AUDIT-C: Alcohol Use Disorders Identification Test–Consumption
BAR: broken adaptive ridge
BMI: body mass index
CAP: controlled attenuation parameter
CI: confidence interval
CT: computed tomography
FIB-4: fibrosis-4 index
GGT: gamma-glutamyl transferase
GLP-1: glucagon-like peptide-1
HALO-MASLD: Health Assessments and Longitudinal Outcomes in MASLD
HbA1c: glycated hemoglobin
HDL: high-density lipoprotein
INR: international normalized ratio
LDL: low-density lipoprotein
LIFT-MASLD: Liver Integrated Forecasting Tool for MASLD
LSM: liver stiffness measurement
MASLD: metabolic dysfunction-associated steatotic liver disease
MASH: metabolic dysfunction-associated steatohepatitis
MetALD: metabolic dysfunction and alcohol-associated liver disease
MPS: microphysiological systems
MRE: magnetic resonance elastography
MRI: magnetic resonance imaging
NAFLD: nonalcoholic fatty liver disease
NCI: National Cancer Institute
NIDDK: National Institute of Diabetes and Digestive and Kidney Diseases
NIH: National Institutes of Health
NRMSE: normalized root mean squared error
PROMIS: Patient-Reported Outcomes Measurement Information System
SCORE: Sparse Canonical Outcome REgression
SD: standard deviation
UPMC: University of Pittsburgh Medical Center

## Appendix

